# Angular reproduction numbers improve estimates of transmissibility when disease generation times are misspecified or time-varying

**DOI:** 10.1101/2022.10.19.22281255

**Authors:** Kris V Parag, Benjamin J Cowling, Ben C Lambert

## Abstract

We introduce the *angular reproduction number* Ω, which measures time-varying changes in epidemic transmissibility resulting from variations in both the effective reproduction number *R*, and generation time distribution *w*. Predominant approaches for tracking pathogen spread either infer *R* or the epidemic growth rate *r*. However, *R* is biased by mismatches between the assumed and true *w*, while *r* is difficult to interpret in terms of the individual-level branching process underpinning transmission. *R* and *r* may also disagree on the relative transmissibility of epidemics or variants (i.e., *r*_*A*_>*r*_*B*_ does not imply *R*_*A*_>*R*_*B*_ for variants A and B). We find that Ω responds meaningfully to mismatches and time-variations in *w* while mostly maintaining the interpretability of *R*. We prove that Ω>1 implies *R*>1 and that Ω agrees with *r* on the relative transmissibility of pathogens. Estimating Ω is no more difficult than inferring *R*, uses existing software, and requires no generation time measurements. These advantages come at the expense of selecting one free parameter. We propose Ω as complementary statistic to *R* and *r* that improves transmissibility estimates when *w* is misspecified or time-varying and better reflects the impact of interventions, when those interventions concurrently change *R* and *w* or alter the relative risk of co-circulating pathogens.

## Introduction

Estimating the rate of spread or transmissibility of an infectious disease is a fundamental and ongoing challenge in epidemiology [1]. Identifying salient changes in pathogen transmissibility can contribute important information to policymaking, providing useful warnings of resurgent epidemics, assessments of the efficacy of interventions and signals about the emergence of new variants of concern [1–3]. The effective or instantaneous reproduction number, *R*, and time-varying growth rate, *r*, are commonly used to characterise pathogen transmissibility. The former statistic is an estimate of the average number of new infections per active (circulating) past infection, while the latter describes the exponential rate of new infection accumulation [4].

Although *R* and *r* are important and popular means of tracking the dynamics of epidemics, they suffer from key limitations that diminish their fidelity and interpretability. Specifically, the meaningfulness of *R* depends on our ability to measure the generation time distribution of the infection under study, *w*. This distribution captures the inter-event times among primary and secondary infections [5] and is convolved with the past infections to define Λ, the time-varying total infectiousness of the disease. The total infectiousness serves as the denominator when inferring *R*, which is the ratio of new infections to Λ. We illustrate all key notation in **Figure 1**. However, infection times and hence *w* are difficult to measure, requiring detailed transmission chain data from contact tracing or transmission studies [6]. Even if these data are available, the estimated *w* (and hence Λ) depends on how inter-event times are sampled or interpreted (e.g., there are forward, backward, intrinsic and realised generation intervals) [7,8].

**Figure 1:**
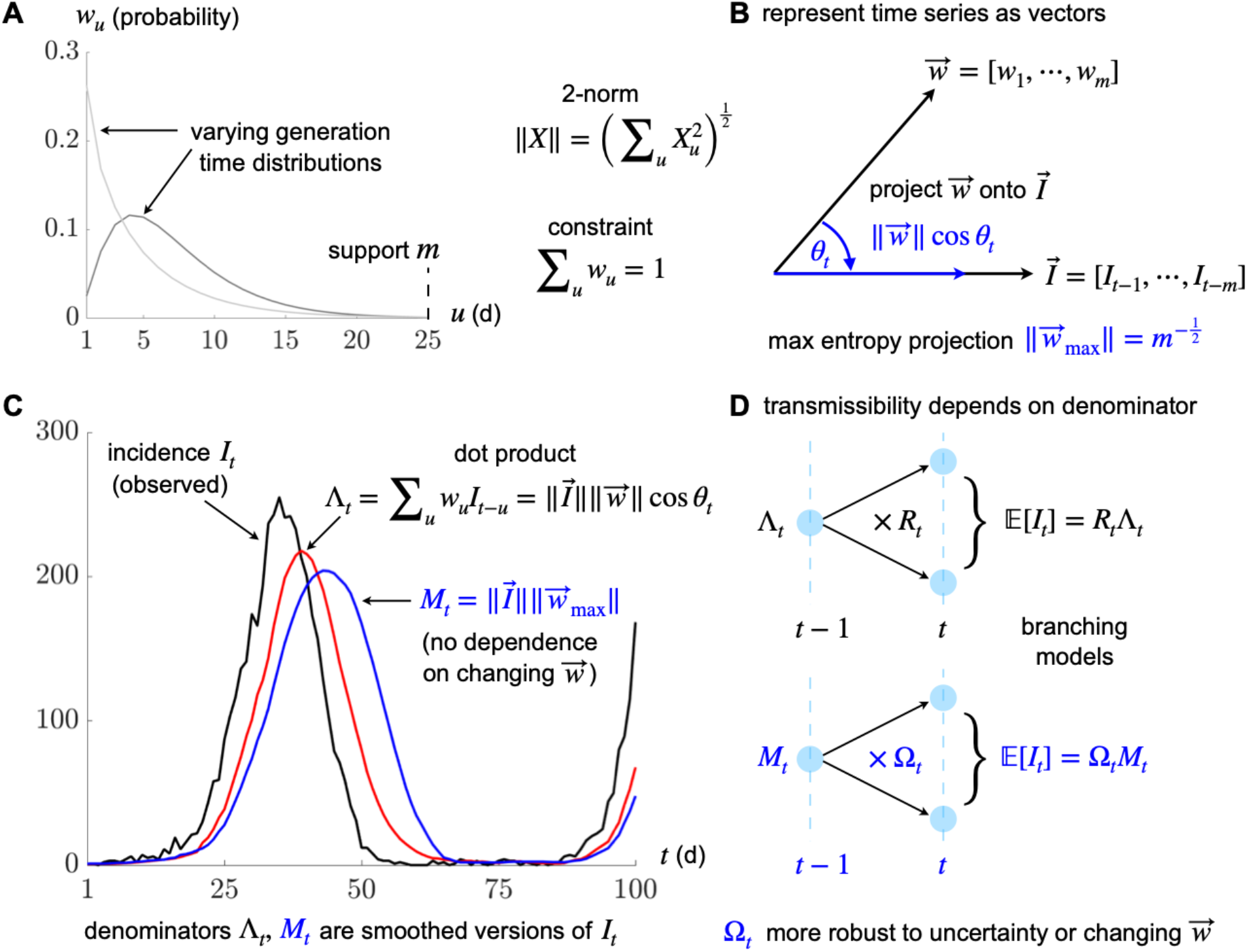
Definitions of transmissibility metrics. Panel A plots generation time distributions that define how past infections cause later ones via the probabilities or weights *w* with support *m*. This involves convolving these weights with past infection incidence *I*. We show in panel B that if we represent *w* and *I* as vectors then the convolution is equivalent to a projection of *w* onto the vector of *I*. Panels C-D illustrate that standard reproduction numbers *R* implicitly apply this projection to compute the denominator Λ. This projection and hence Λ is sensitive to the *w*, meaning that if the distribution switches between the two from panel A, our estimates of *R* become biased (often changes in generation time are difficult to measure). Our new metric Ω maximises the projection from panel B to reduce sensitivity (practically this involves a window based on *m*) leading to a new denominator *M* in panel C. This maintains the branching process interpretation of the epidemic in panel D, while improving transmissibility estimate robustness.

Workarounds, such as approximating *w* by the serial interval distribution [9], which describes inter-event times between the onset of symptoms, or inferring *w* from this distribution [10], do exist but suffer from related problems [6]. Consequently, *w* and Λ can often be misspecified, biasing *R* and likely misrepresenting the true branching process dynamics of epidemics. While *r* is more robust to *w* misspecification (it only depends on the log gradient of the smoothed infection time series) [4], it lacks the individual-level informativeness and interpretation of *R*. Given estimates of *r*, it is unclear how to derive the proportion of new infections that need to be suppressed (roughly *R*^*-1*^), herd immunity thresholds (related to 1-*R*^*-1*^) or the probability of epidemic elimination and establishment (both linked to *R*^*-N*^ for *N* infections) [11–13]. The only known means of attaining such information converts *r* into *R* using estimates of *w* [14].

Difficulties in accurately inferring generation times therefore cause practical bottlenecks that constrain our ability to measure pathogen transmissibility. These problems are worsened as recent studies have empirically found that generation times also vary substantially with time (i.e., *w* is non-stationary) [15]. These variations may correspond to different epidemic phases [16], emerging variants of concern [17] and coincide with the implementation of interventions [18]. These are precisely the situations in which we also want to infer *R*. However, concurrent changes in *R* and *w* are rarely identifiable, and *r* inextricably groups the effects of *w* and *R* on transmissibility. While high quality, longitudinal contact tracing data [19] can potentially resolve these identifiability issues, this is an expensive and logistically hard solution. Here we propose another means of alleviating the above problems and complementing the insights provided by *R* and *r* – the *angular reproduction number*, Ω.

The angular reproduction number defines transmissibility as a ratio of new infections to *M*, the root mean square number of past infections over a user-defined window δ. Because it replaces Λwith *M*, a quantity that does not require knowledge of generation times, Ω is more robust to the problems of inferring *w*. We demonstrate that Ω is able to measure the overall changes in transmissibility caused by fluctuations in both *R* and *w*. Moreover, we prove that Ω has similar threshold properties to *R*, maintains much of its individual-level interpretation and is a useful metric for communicating transmissibility. This last point follows as we only need to quote Ω and the known window δ to generalise our estimates of transmissibility to different settings. In contrast, the meaningfulness of *R* is contingent on the unknown or uncertain *w*. Downstream studies sometimes use *R* outside of its generation time context [20], while dashboards aiming at situational awareness commonly quote *R* without *w*, introducing biases and interpretability problems into how disease spread is communicated [21].

Additionally, we demonstrate how *r* and *R* can easily disagree on relative transmissibility, both across time and for co-circulating variants. Unmeasured changes in *w* over time can cause *R* and *r* to vary in opposite directions (one signals an increase in transmissibility and the other a decrease). Similarly, co-circulating pathogens with different but stationary and known *w*, may possess contradictory *R* and *r* value rankings i.e., for variants A and B, *r*_*A*_ > *r*_*B*_ does not imply *R*_*A*_ > *R*_*B*_. These issues are amplified when interventions (which can change *w, R* or both [18]) occur, obscuring notions of the relative risk of spread. However, we find *r*_*A*_ > *r*_*B*_ guarantees Ω_*A*_ > Ω_*B*_ and that Ω is consistent with *r* across time even when *w* changes.

Last, while we may also convert *r* into threshold statistics about 1 by using a free parameter together with a transformation from [14], we show that Ω is more robust to choices of its free parameter than those statistics, which implicitly make stronger assumptions (**Supplementary Information**). These robustness and consistency properties of Ω reinforce its usefulness for tracking and comparing outbreak spread and emerge from its maximum entropy approach to managing uncertain generation time distributions. We propose Ω as a complementary statistic that can be integrated with *R* and *r* to present a more comprehensive perspective on epidemic transmissibility, especially when *w* is poorly specified or varying with time.

## Results

### Angular reproduction numbers

The epidemic *renewal model* [22] provides a general and flexible representation of disease transmission. It defines how the incidence of new infections at time *t*, denoted *I*_*t*_, depends on the effective or *instantaneous reproduction number, R*_*t*_, and the past incident time series of infections, 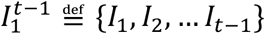. This results in the conditional moment relationship in **Eq. (1)** [9]. Generally, we use 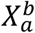 to denote the time series {*X*_*a*_, *X*_*a*+1_, …, *X*_*b*−1_, *X*_*b*_ } and **E**[*X*|*Y*] for the expectation of *X* over possible epidemic trajectories given known variables *Y*. Where obvious, and for convenience, we sometimes drop *Y* in **E**[*X*|*Y*], writing **E**[*X*].

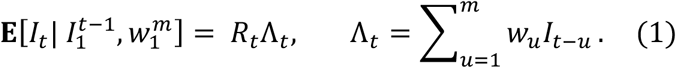

In this model Λ_*t*_ is known as the *total infectiousness* and summarises the weighted influence of past infections. The set of weights *w*_*t*_ for all *u* defines the *generation time distribution* of the infectious disease with 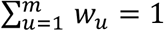, and *m* as the support of this distribution, which we assume to be practically finite [14]. When the time series is shorter than *m* we truncate and renormalise the *w*_*u*_. Commonly, the stochasticity around the expectation *R*_*t*_Λ_*t*_ is modelled using either Poisson or negative binomial count distributions [1,12].

Although **Eq. (1)** has successfully been applied to model many diseases including COVID-19, Ebola virus disease, pandemic influenza and measles, among others, it has one major flaw – it assumes that the generation time distribution is fixed or stationary and known [9]. If this assumption holds (we ignore surveillance biases [9,23] until the Discussion), **Eq. (1)** allows epidemic transmissibility to be summarised by fluctuations of the time-varying *R*_*t*_ parameters. This follows because the sign of *R*_*t*_ − 1 determines if *I*_*t*_ will increase or decline relative to the total infectiousness Λ_*t*_. This reproduction number can be linked to the *instantaneous epidemic growth rate, r*_*t*_, using the moment generating function of the generation time distribution [14].

Consequently, from *R*_*t*_, we obtain temporal information about the rate of pathogen spread and its mechanism i.e., we learn how many new infections we can expect per circulating infection because 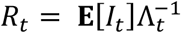. As *R*_*t*_ is a threshold parameter, we know that we must block at least a fraction 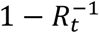 of new infections to suppress epidemic growth (*R*_*t*_ = 1 signifies that *r*_*t*_ = 0). The time scale over which this suppression is achievable [14] and our ability to detect these changes in *R*_*t*_ [24] in the first place, however, are determined by the generation times.

Recent works emphasise that the assumption of a known or fixed generation time distribution is often untenable, with appreciable fluctuations caused by interventions [15,18] and emerging pathogenic variants [17] or occurring as the epidemic progresses through various stages of its lifetime [5]. Substantial biases in *R*_*t*_ can result (because its denominator Λ_*t*_ is incorrectly specified [4]), which even impede optimal Bayesian inference algorithms [25]. As *R*_*t*_ is the predominant metric of transmissibility, contributing key evidence towards infectious disease policymaking [1], this may potentially obscure situational awareness or misinform intervention planning. While improved and intensive contact tracing can provide updated generation time information, this is usually difficult and expensive. We propose a robust alternative.

We redefine Λ_*t*_ by recognising it as a dot product between the vectors of generation time probabilities 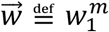 and the past incidence 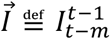 over the support of the generation time distribution, *m*. This gives the left equality of **Eq. (2)** with the Euclidian norm of 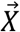 as 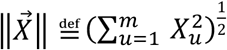 and *θ*_*t*_ as the time-varying angle between 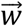 and 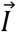. This equality holds for non-stationary generation times i.e., both 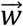 and 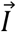 can have elements that change over time. We illustrate this notation and elements of the subsequent derivation in **Figure 1. Eq. (2)** implies that the count of new infections (for any *R*_*t*_) is maximised when *θ*_*t*_ is minimised i.e., when the temporal profile of past infections matches the shape of the generation time distribution.

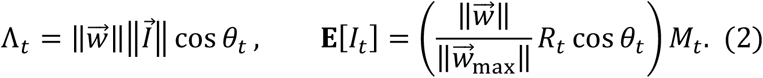

We can compute the root mean square of the incidence across the support of the generation time distribution as 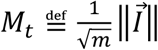. Under the constraint that 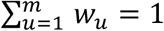 (if *t* − 1 < *m* we truncate this distribution to sum to 1 – this is an edge effect of the epidemic) then the maximum possible value of the generation time norm is 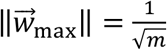. This is achieved by the maximum entropy generation time distribution of 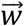, which is uniform (has *m* entries of 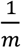).

Combining these definitions with **Eq. (1)**, we derive the second expression in **Eq. (2)** for the expected number of new infections at time *t*. This may seem an unnecessarily complicated manipulation of the standard renewal model, but it admits a novel and important insight – we can separate the influences of the reproduction numbers and the generation time distribution (together with its changes) on epidemic transmissibility. These multiply *M*_*t*_, which defines a new denominator – the root mean square number of past infections (this is also the average signal power of the past infection time series) – that replaces the total infectiousness Λ_*t*_.

Consequently, we define a new metric in **Eq. (3)**, the *angular reproduction number* Ω_*t*_, which multiplies *R*_*t*_ by the scaled projection of the generation time distribution, 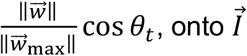, the past incidence vector (see **Figure 1**). This means that Ω_*t*_ is a time-varying ratio between the expected infection incidence and the past root mean square incidence *M*_*t*_. We use the term reproduction number for Ω_*t*_ due to its relation to *R*_*t*_, the similarity of **Eq. (2)** and **Eq. (3)** and because of its threshold properties, which we explore in the next section.

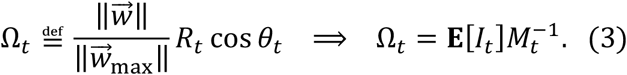

This metric captures all possible variations that impact the ability of the epidemic to transmit. It responds to both changes in *R*_*t*_ and the generation time distribution. The latter would scale 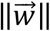 and rotate *θ*_*t*_, which is why we term this angular. The benefit of compactly describing both types of transmissibility changes does come with a trade-off in interpretability as it may be harder to intuit the meaning behind **E**[*I*_*t*_] = Ω_*t*_*M*_*t*_ than the more usual **E**[*I*_*t*_] = *R*_*t*_Λ_*t*_.

We argue that this is not the case practically because Λ_*t*_ is frequently misspecified [15,26], obscuring the meaning of *R*_*t*_. In contrast, *M*_*t*_ does not depend on generation time assumptions (beyond characterising its support *m*). We remove structural uncertainty induced by the often unknown *w*_*u*_ because *M*_*t*_ is a maximum entropy version of Λ_*t*_ i.e., 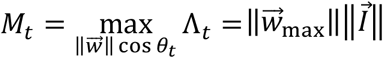 subject to 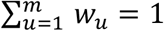. We also find that *M*_*t*_ = Λ_*t*_ and hence Ω_*t*_ = *R*_*t*_, when the past incidence is flat (as then Λ_*t*_ = *M*_*t*_ and *w*_*u*_ has no effect). This defines the important and universal equilibrium condition Ω_*t*_ = *R*_*t*_ = 1. There is further convergence for branching process models [27] with timesteps at its fixed generation time, as then trivially *w*_1_ = 1.

### Relationship to popular transmissibility metrics

Having defined the angular reproduction number above, we explore its properties and show why it is an interesting and viable measure of transmissibility. We examine an exponentially growing epidemic with incidence *I*_*t*_ = *I*_0_*e*^*rt*^ and constant growth rate *r*. This model matches the dynamics of fundamental *compartmental models* such as the SIR and SEIR (in the limit of an excess of susceptible individuals) and admits the known relation *gr* = (*R* − 1) [28], with *g* as the mean generation time. We assume growth occurs over some period of δ and compute Ω_*t*_ as the ratio 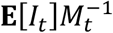 from **Eq. (3)**. Since this model is deterministic **E**[*I*_*t*_] = *I*_*t*_ = *I*_0_*e*^*rt*^.

We evaluate *M*_*t*_ from its definition above as 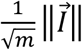 with δ = *m* and using the continuous-time expression for 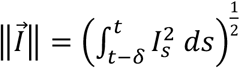. This yields 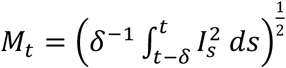 with 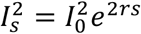 from the exponential incidence equation and evaluates to 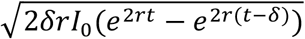. Substituting this into 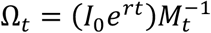 results in the left relation in **Eq. (4)**.

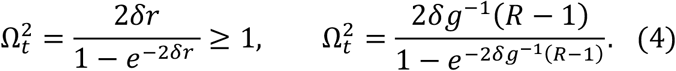

Several important points follow. First, as *x* ≥ 1 − *e*^−*x*^ for every *x* ≥ 0, then Ω_*t*_ − 1 and *r* are positive too (an analogous argument proves the negative case). Second, we substitute for *r* using the compartmental *R-r* relationship *gr* = (*R* − 1) to get the right-side relation of **Eq. (4)**. Applying L’ Hopital’s rule we find 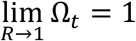. We hence confirm the threshold behaviour of Ω_*t*_ i.e., the sign of Ω_*t*_ − 1 and *R*_*t*_ − 1 are always consistent (for all values of δ > 0).

Third, we see that constant growth rates imply constant angular reproduction numbers. The converse is also true, and we may input time-varying growth rates, *r*_*t*_, into **Eq. (4)** to estimate Ω_*t*_. These properties hold for any δ, which is now a piecewise-constant time window. We plot the ramifications of **Eq. (4)** in **Figure 2**. Further, in **Table *1*** we summarise how Ω_*t*_ relates to predominant *R*_*t*_ and *r*_*t*_ metrics. We explore some properties in this table in later sections (in addition to reinforcing our analyses with stochastic models) and demonstrate that relationships among *r*_*t*_, *R*_*t*_ and Ω_*t*_ have important consequences when comparing outbreaks subject to interventions and variations in generation times.

**Table 1:** Summary of transmissibility metrics. We list important relationships among the instantaneous growth rate (*r*), the instantaneous or effective reproduction number (*R*) and the angular reproduction number (Ω) and assess their value as measures of transmissibility.

| Metric property | Growth $r$ | Effective $R$ | Angular $\Omega$ |
| --- | --- | --- | --- |
| Definition of transmissibility | $r_t \stackrel{\text{def}}{=} \frac{d \log \mathbf{E}[I_t]}{dt}$ | $R_t \stackrel{\text{def}}{=} \frac{\mathbf{E}[I_t]}{\Lambda_t}$ | $\Omega_t \stackrel{\text{def}}{=} \frac{\mathbf{E}[I_t]}{M_t}$ |
| Pathogen spread threshold | $r_t > 0$ | $R_t > 1$ | $\Omega_t > 1$ |
| Biased by generation time $\vec{w}$ assumed, given curve $I_1^t$ | Insensitive to the assumed $\vec{w}$ | Biased when $\vec{w}$ is misspecified | Signals changes in $\vec{w}$ and $R_t$ |
| Ranking risk of outbreaks or variants by spreading rate | $r_A > r_B \Rightarrow$ variant $A$ spreads faster | $r_A > r_B \nRightarrow R_A > R_B$ (inconsistent) | $r_A > r_B \Rightarrow \Omega_A > \Omega_B$ (consistent) |
| Short-term predictive power | Negligible differences among metrics in prediction quality |  |  |
| Non-dimensional metric | No, inverse time | Yes, both have no units, scalable |  |
| Individual-level interpretability | Not obvious | New infections per circulating ones |  |
| Computability if $\vec{w}$ unknown | Yes (smooth $I_1^t$) | Not possible | Yes, for any $\delta$ |

**Figure 2:**
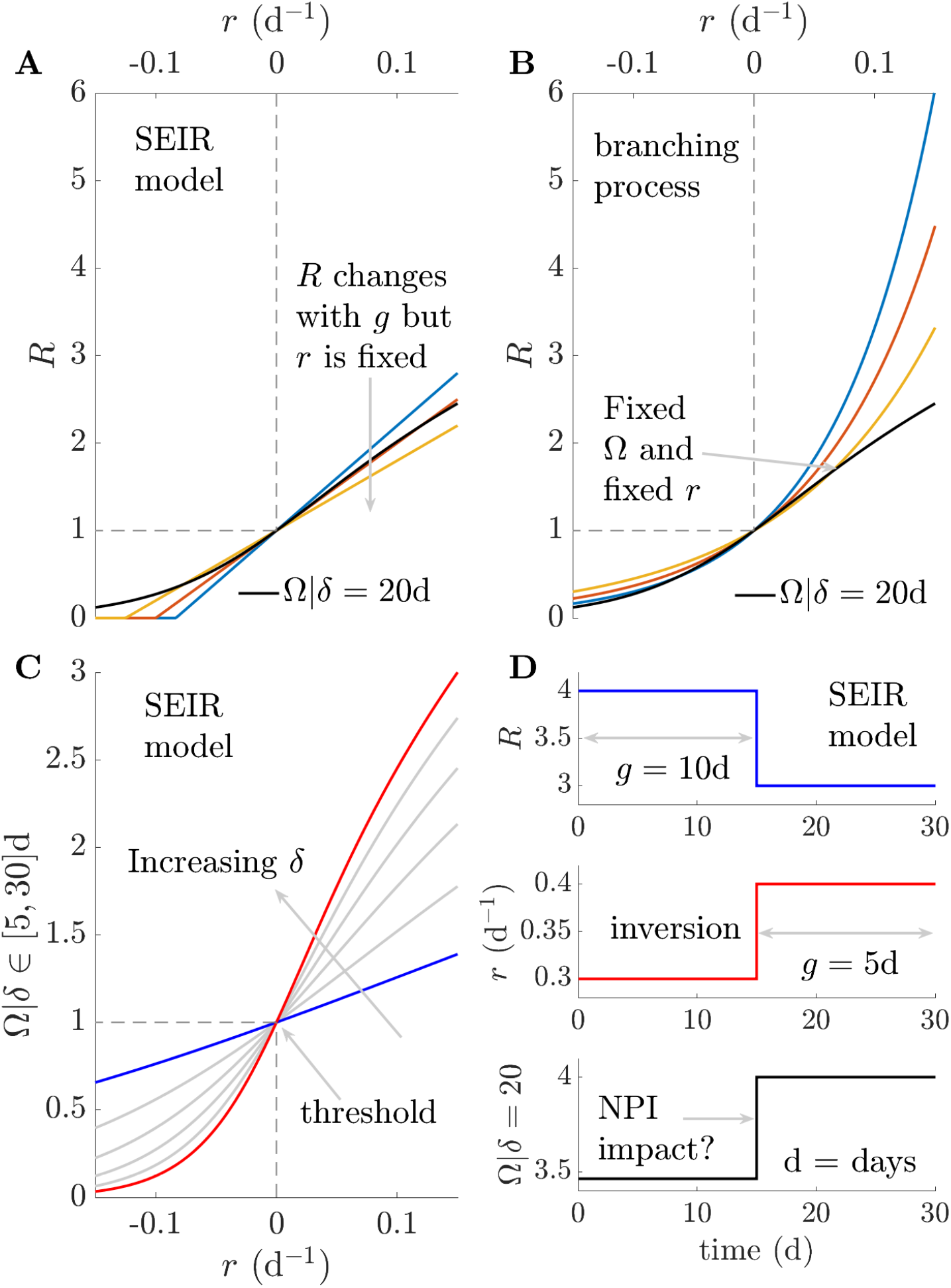
Relationships among transmissibility metrics. Panel A and B show how growth rates (*r*) and reproduction numbers (*R*) have diverse functional relationships (see [14,30]) for SEIR models with an excess of susceptible individuals and branching processes. Coloured lines indicate *R* at different mean generation times (*g*). Black lines highlight a single functional relationship between angular reproduction numbers Ω and *r* at all *g*, using a window *8* of 20d. Panel C shows that while Ω varies with choice of δ (increasing from blue to red and computed from **Eq. (4)**), we have a bijective relationship with *r*. Panel D indicates that *R* and *r* can signify inverted changes e.g., an NPI reducing *R* and *g* may increase *r*, raising questions about impact (see [15,18]). Here Ω converts *r* into a consistent transmissibility metric (also from **Eq. (4)**).

Note that we may also invert the relationship in **Eq. (4)** to estimate *r*_*t*_ from Ω_*t*_ (see Methods for details). This involves solving **Eq. (5)**, where *W*_*k*_(*x*) is the Lambert W function with index *k* ∈ [0, −1] (this range results from the indicator **1**(y)) [29].

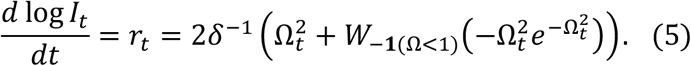

A central implication of **Eq. (4)** and **Eq. (5)** is that we can infer angular reproduction numbers directly from growth rates or vice versa, without requiring knowledge of the generation times.

We further comment on connections between angular and effective reproduction numbers using a deterministic *branching process* model, which is also foundational in epidemiology. We again focus on growth, which is geometric as this is a discrete-time process with time steps scaled in multiples of the mean generation time *g*. Here incidence is *I*_*t*_ = *R*^*t*^ and 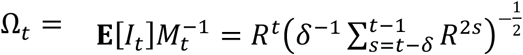, with window δ in units of *g*. If δ = 1 we recover Ω_*t*_ = *R*. If *R* = 1, then Ω_*t*_ = *R* for all δ. For growing epidemics, as δ increases, Ω_*t*_ > *R* because we reference present incidence to smaller past infections (or denominators). The opposite occurs if the epidemic declines. This may seem undesirable, but we argue that Ω_*t*_ improves overall practical transmissibility measurement because *g* will likely be misspecified or vary with time.

Any *g* mismatches bias *R*, limiting its interpretation, meaningfulness and making comparisons among outbreaks or pathogenic variants difficult, because we cannot be certain that our denominators correspond. This is particularly problematic when estimates of *R* obtained from a modelling study are incorporated as parameters into downstream studies without accounting for the generation time context on which those estimates depend. However, by additionally communicating Ω and δ, we are sure that denominators match and that we properly include the influences of any *g* mismatches. Choosing δ is also no worse (and more explicit) than equivalent window assumptions made when inferring *R* and *r* [4,30] In the **Supplementary Information** we perform analyses of window choices for Ω and other threshold metrics.

Last, we illustrate how Ω_*t*_ relates to other key indicators of epidemic dynamics such as herd immunity and elimination probabilities. As our derivation replaces **Eq. (1)** with 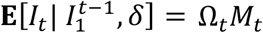 for the same observed incidence, these indicators are also readily obtained. Assuming Poisson noise, the elimination probability 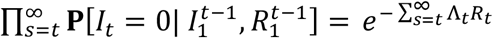 is replaced by 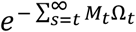, and has analogous properties [31]. Herd immunity, which traditionally occurs when a fraction 1 − *R*^−1^ of the population is immune is approximated by 1 − Ω^−1^ (since both metrics possess the same threshold behaviour) [11]. In a subsequent section we demonstrate that one-step-ahead incidence predictions from both approaches are also comparable.

### Responding to variations in generation time distributions

We demonstrate the practical benefits of Ω_*t*_ using simulated epidemics with non-stationary or time-varying generation time distributions. Such changes lead to misspecification of Λ_*t*_ in **Eq. (1)**, making estimates of the effective reproduction number *R*_*t*_, denoted 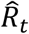, a poor reflection of the true underlying *R*_*t*_. In contrast variations in the estimated 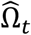 are a feature (see **Eq. (3)**) and not a bug (for some chosen δ we control *M*_*t*_, which is not misspecified). We simulate epidemics with Ebola virus or COVID-19 generation times from [32,33] using renewal models with Poisson noise [9]. We estimate both the time-varying *R*_*t*_ and Ω_*t*_ using *EpiFilter* [25], which applies Bayesian algorithms that minimise mean square estimation error.

Inferring Ω_*t*_ from incident infections, 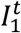, requires only that we replace the input Λ_*t*_ with *M*_*t*_ in the estimation function and that we choose a window δ for computing *M*_*t*_. We provide software for general estimation of Ω_*t*_ and code for reproducing this and all other analyses in this paper at https://github.com/kpzoo/Omega. We heuristically set δ ≈ 2*g*_0_ as our window with *g*_0_ as the original mean generation time of each disease from [32,33]. We find (numerically) that this δ ensures 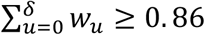 over many possible gamma distributed generation times i.e., it is long enough to cover most of the likely probability mass of unknown changes to the generation time distributions, which cause time-varying means *g*_*t*_. In general, we find that an overly small δ tends to neglect important dynamics, while too large a δ induces edge effects. The Methods and **Supplementary Information** provide for more information on choosing δ.

Our results are plotted in **Figure 3**. We show that 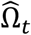 responds as expected to both changes in the true *R*_*t*_ and 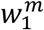 subject to the limits on what can be inferred [24]. In **Figure 3** we achieve changes in 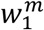 by altering the mean generation time *g*_*t*_ by ratios that are similar in size to those reported from empirical data [15]. In contrast, we observe that 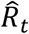 provides incorrect and overconfident transmissibility estimates, which emerge because its temporal fluctuations also have to encode structural differences due to the misspecification of 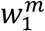. These can strongly mislead our interpretation and understanding of the risk posed by a pathogen.

**Figure 3:**
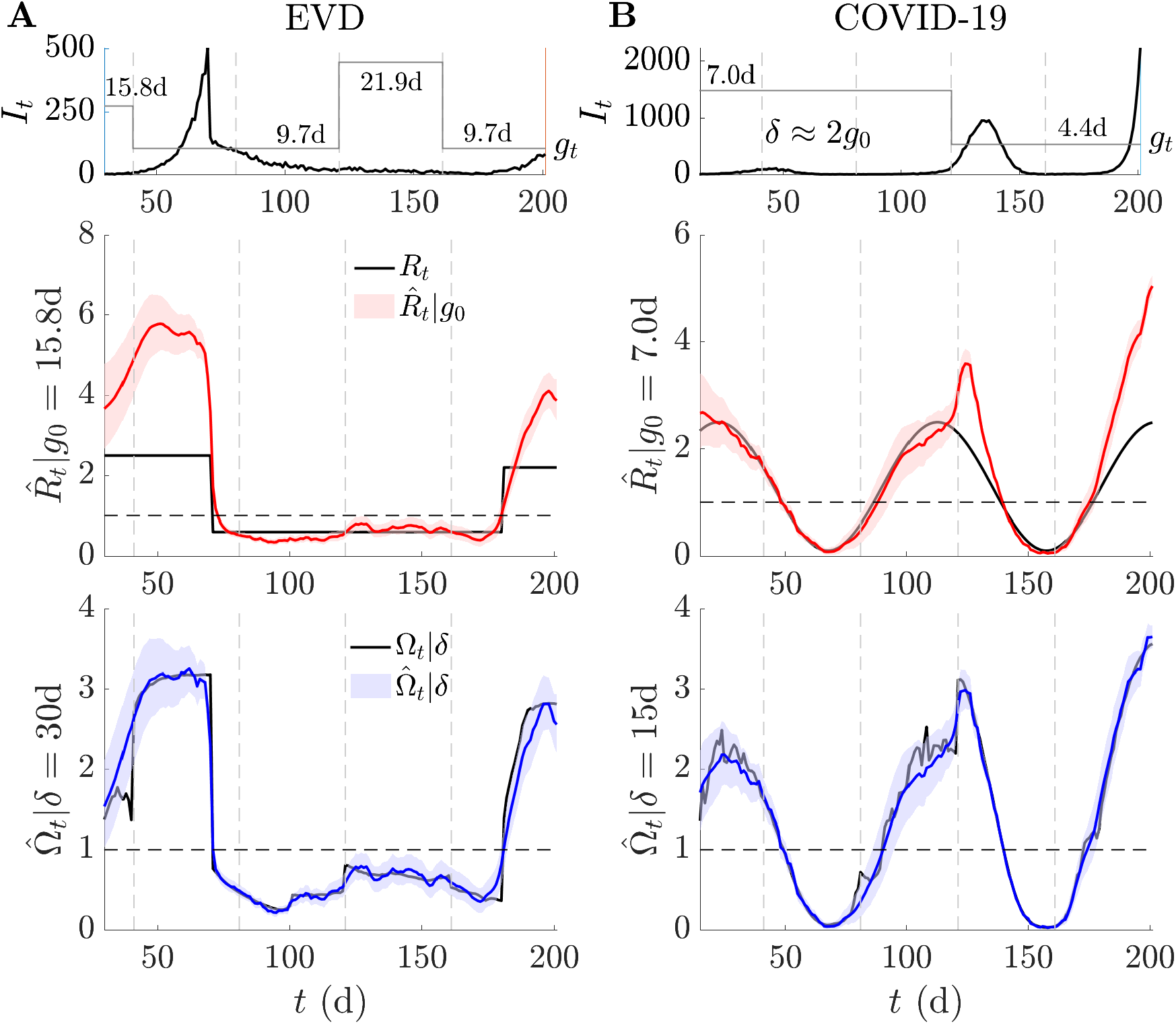
Estimating transmissibility under temporal variations in generation times. We simulate epidemic incidence curves (black) using generation time distributions of Ebola virus disease (EVD) [32] and COVID-19 [33] in panels A and B. The means of these distributions (*g*) vary over time (grey piecewise, starting from original mean *g*_*0*_), but we fix their variance at their original values. We find substantial bias in *R* estimated from the initial EVD and COVID-19 generation times (red with 95% credible intervals, true value in black). These estimates try to compensate for generation time mismatches and changes in an uncontrolled manner that obscures interpretation. However, Ω responds as we expect (blue with 95% credible intervals, window δ, true value in black) and we infer change-points due to both *R* and *g* fluctuations (subject to bounds induced by noise i.e., at low incidence inference is more difficult [24]). Our estimates derive from EpiFilter [25] with default settings and we truncate time series to start from δ to remove any edge effects. Vertical dashed lines highlight times at which we change *g* or keep it fixed. When it is fixed, Ω infers no spurious changes.

We can derive alternative threshold statistics that relate to *r*_*t*_ and do not explicitly depend on the generation time by applying monotonic transformations from [14]. In theory these should have comparable behaviour around the critical point of 1 to both *R*_*t*_ and Ω_*t*_. We investigate these statistics in the **Supplementary Information**, computing them across the simulations of **Figure 3**. We find that they require stronger assumptions than Ω_*t*_ (i.e., they fix distributional formulae for generation times), possess at least as many free parameters as Ω_*t*_ and are less robust to changes in those parameters (often strongly over-estimating transmissibility), than Ω_*t*_ is to fluctuations in δ. This confirms that angular reproduction numbers can complement standard metrics, improving transmissibility estimates when generation times are changing, or unknown and forming part of a more comprehensive suite of outbreak diagnostics.

### Ranking epidemics or variants by transmissibility

Misspecification of generation time distributions, and corresponding misestimation of *R* as in **Figure 3**, also plays a crucial role when assessing the relative transmissibility of pathogens, variants of concern or even outbreaks (where we may want to contrast the spread of contagion among key demographic or spatial groups). As shown in **Figure 2**, these variations can mean that increases in the growth rate *r*_*t*_ actually signify decreases in the effective reproduction number *R*_*t*_ or that a pathogen with a larger *r*_*t*_ can have a smaller *R*_*t*_. Here we illustrate that these issues can persist even if the generation time distributions of pathogens are correctly specified and remain static, obscuring our understanding of relative transmission risk.

In **Figure 4** we simulate epidemics under two hypothetical variants of two pathogens. We use EVD and COVID-19 generation time distributions from [32,33] to define our respective base variants. For both pathogens we specify the other variant by reducing the mean generation of each base but fixing the variance of the generation times. Reductions of this type are plausible and have been measured for COVID-19 variants [17]. All 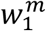 distributions are stationary and known in this analysis. We discover that changes in *R*_*t*_ alone can initiate inversions in the relative growth rate of different variants or epidemics. As far as we can tell, this phenomenon has not been explicitly investigated. Given that interventions can change *R*_*t*_ in isolation or in combination with 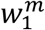 [15,18], this effect has the potential to be widespread. We determine the mathematical conditions for this inversion in the **Supplementary Information**.

**Figure 4:**
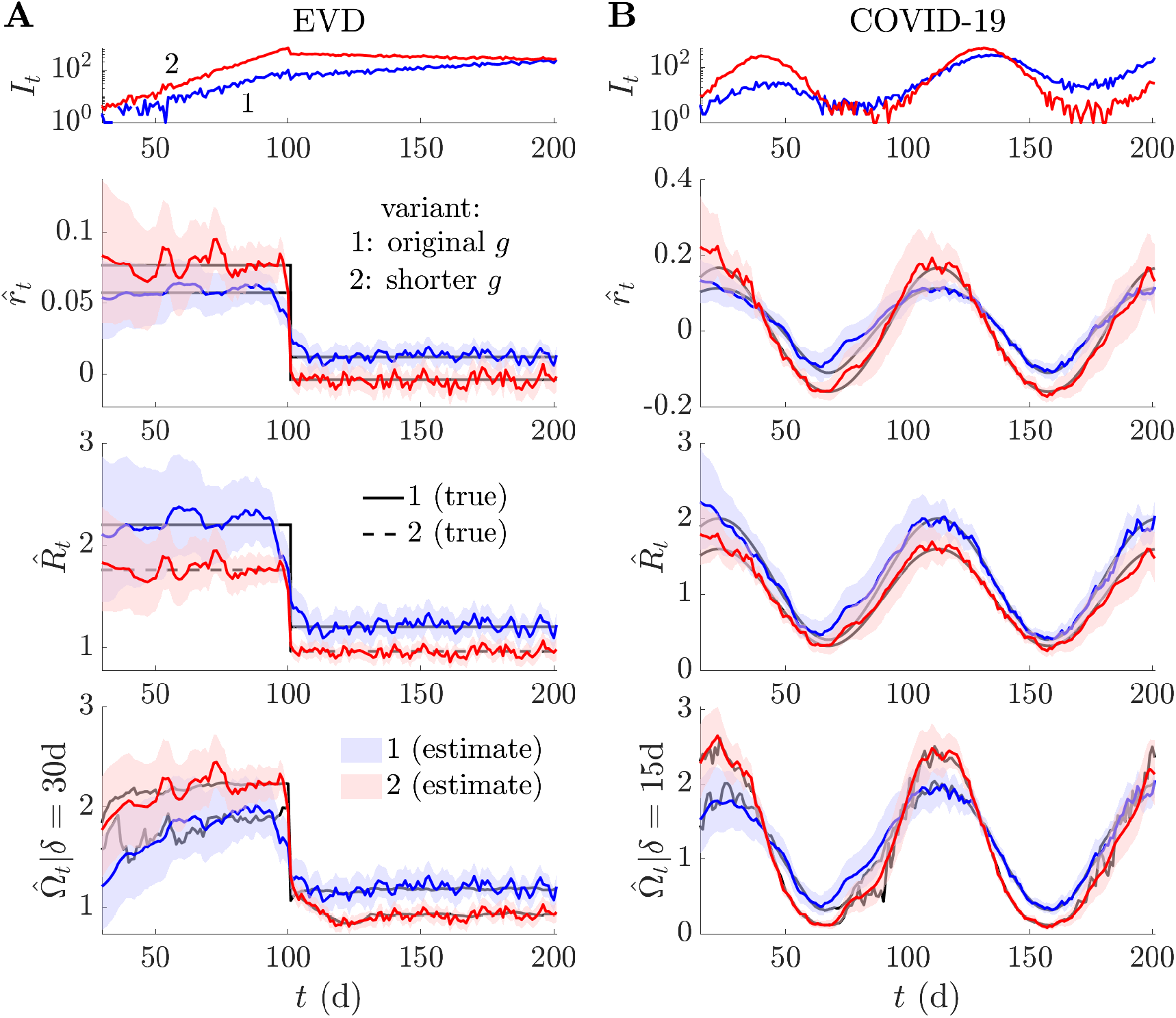
Comparing transmissibility across outbreaks, variants or even diseases. We simulate epidemics of variant 1 in blue (with estimates of metrics also in blue) under standard generation time distributions of Ebola virus disease (EVD) [32] and COVID-19 [33] in panels A and B. In red (with estimates also in red) we overlay simulations in which the generation time of these diseases is 40% and 50% shorter (than the blue epidemics), which may indicate a new co-circulating variant 2 or another epidemic with different properties (e.g., in a higher risk group). We demonstrate (for the first time to our knowledge) that changes in *R* due to an intervention (or release of one) may invert the relative growth rates (*r*) of the epidemics (see **Supplementary Information** for mathematical intuition for this inversion). The mismatches in the *R*-*r* rankings alter perceptions of relative risk, making transmissibility comparisons difficult. However, Ω classifies the risk of these epidemics in line with their realised growth rates, while still offering the individual-level interpretability of a reproduction number. True values are in black and all estimates (with 95% credible intervals) are outputs from EpiFilter [25] with default settings. We truncate the time series to start from δ to remove any edge effects.

Interestingly, the angular reproduction numbers of **Figure 4** do preserve an ordering that is consistent with growth rates, while maintaining the interpretability (e.g., threshold properties) of reproduction numbers. Hence, we argue that Ω_*t*_ blends advantages from both *R*_*t*_ and *r*_*t*_ [4] and serves as a useful outbreak analytic for understanding and conveying the relative risk of spread of differing pathogens or pathogen strains, or of spread among different spatial and demographic groups. Recent studies have only begun to disentangle component drivers of transmission, including the differing effects that interventions can introduce (e.g., by defining the strength and speed of control measures [34]) and the diverse properties of antigenic variants [17]. We believe that Ω can play a distinctive role in accelerating these investigations.

### Reproduction numbers for explanation or prediction?

We highlight an important but underappreciated subtlety when inferring the transmissibility of epidemics – that the value of accurately estimating *R, r* and Ω largely depends on if our aim is to explain or predict [35] the dynamics of epidemics. The above analyses have focussed on characterising transmissibility to explain mechanisms of spread and design interventions. For these problems, misestimation of parameters, such as *R*, can bias our assessment of outbreak risk and hence misinform the implementation of control measures. An important concurrent problem aims to predict the likely incidence of new infections from these estimates. This involves projecting the epidemic dynamics forward in time to infer upcoming infection patterns.

Here we present evidence that the solution of this problem, at least over short projection time horizons, is robust to misspecification of generation times provided both the incorrect estimate and the misspecified denominator are used in conjunction. We repeat the analyses of **Figure 3** for 200 replicate epidemics and apply EpiFilter [25] to obtain the one-step-ahead predictive distributions 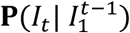 for every *t*. We compute the predicted mean square error (PMSE) and the accumulated predictive error (APE). These scores, which we denote as 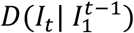, average square errors between mean predictions and true incidence and sum log probabilities of observing the true incidence from the predicted distribution respectively [36,37]. We plot the distributions of scores over replicates and illustrate individual predictions in **Figure 5**.

**Figure 5:**
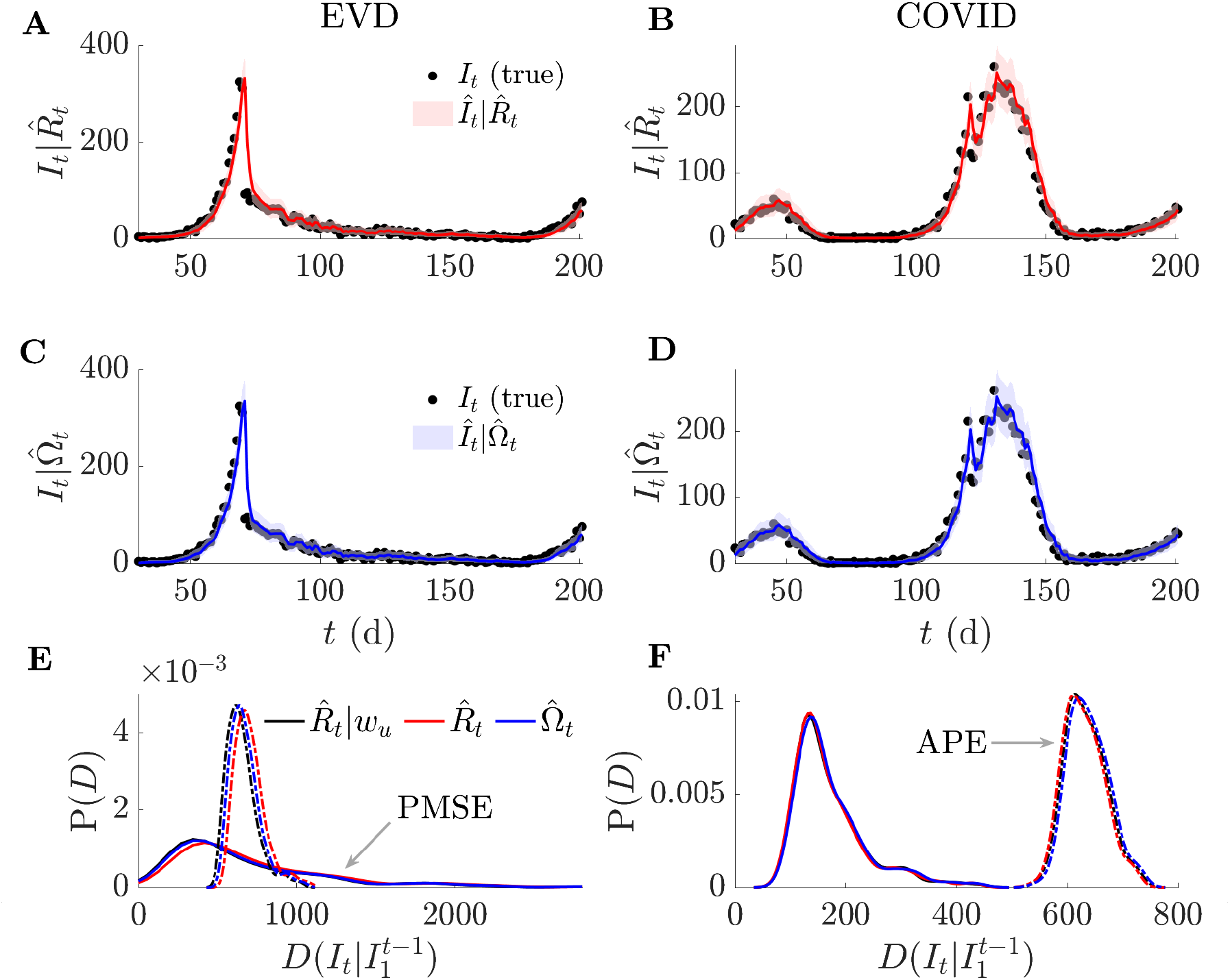
One-step-ahead prediction accuracy and model mismatch. We simulate 200 replicates of the epidemics from **Figure 3**, which involve non-stationary changes to EVD and COVID-19 generation times. We use estimates of effective, *R*, and angular, Ω, reproduction numbers to produce successive one-step-ahead predictions and assess their accuracy to the simulated (true) incidence. Panels A-D provide a representative example of a single simulated epidemic (true incidence shown as black dots) and the *R* and Ω one-step ahead predictions (red and blue respectively with 95% credible intervals). In panels E-F we formally compute accuracy using distance metrics, *D*, based on accumulated prediction errors (APE, dashed) and prediction mean square errors (PMSE, solid) for all 200 replicates from *R*, Ω and *R* given knowledge of the generation time changes i.e., *R*|*w*. We obtain distributions of *D* by applying kernel smoothing. We find negligible differences in predictive power from all approaches.

We find only negligible differences among the one-step-ahead predictive accuracies of the *R* estimated given knowledge of the changing generation times (*R*|*w*), the *R* estimated assuming an unchanged (and hence wrongly specified) *w* and our inferred Ω As APE and PMSE also measure model suitability, their similarity across the three estimates demonstrate that, if the problem of prediction is of interest, then incorrect generation time choices are not important as long as the erroneous denominator (Λ_*t*_) and estimate (*R*_*t*_) are used together. If this estimate is however used outside of the context of its denominator (e.g., if it is simply input into other studies), then inaccurate projections will occur (in addition to poor estimates). As multi-step-ahead predictions can be composed from iterated one-step-ahead ones [38], we conjecture that subtleties between prediction and explanation are likely to also apply on longer horizons.

### Empirical example: COVID-19 in mainland China

We complete our analysis by illustrating the practical usability of Ω on an empirical case study where generation time changes are known to have occurred. In [15], the dynamics of COVID-19 in mainland China are tracked across January and February 2020. Transmission pair data indicated that the serial interval of COVID-19 shortened across this period leading to biases in the inferred *R* if updated serial intervals are not used. Here serial intervals, which measure the lag between the symptom onset times of an infector and infectee are used as a proxy for the generation time. **Figure 6** presents our main results. We find Ω (blue), which requires no serial interval information, behaves similarly to the *R* (red) inferred from the time-changing *w*. Both metrics appear less biased than estimates of *R* (green) that assume a fixed serial interval. This is largely consistent with the original investigation in [15].

**Figure 6:**
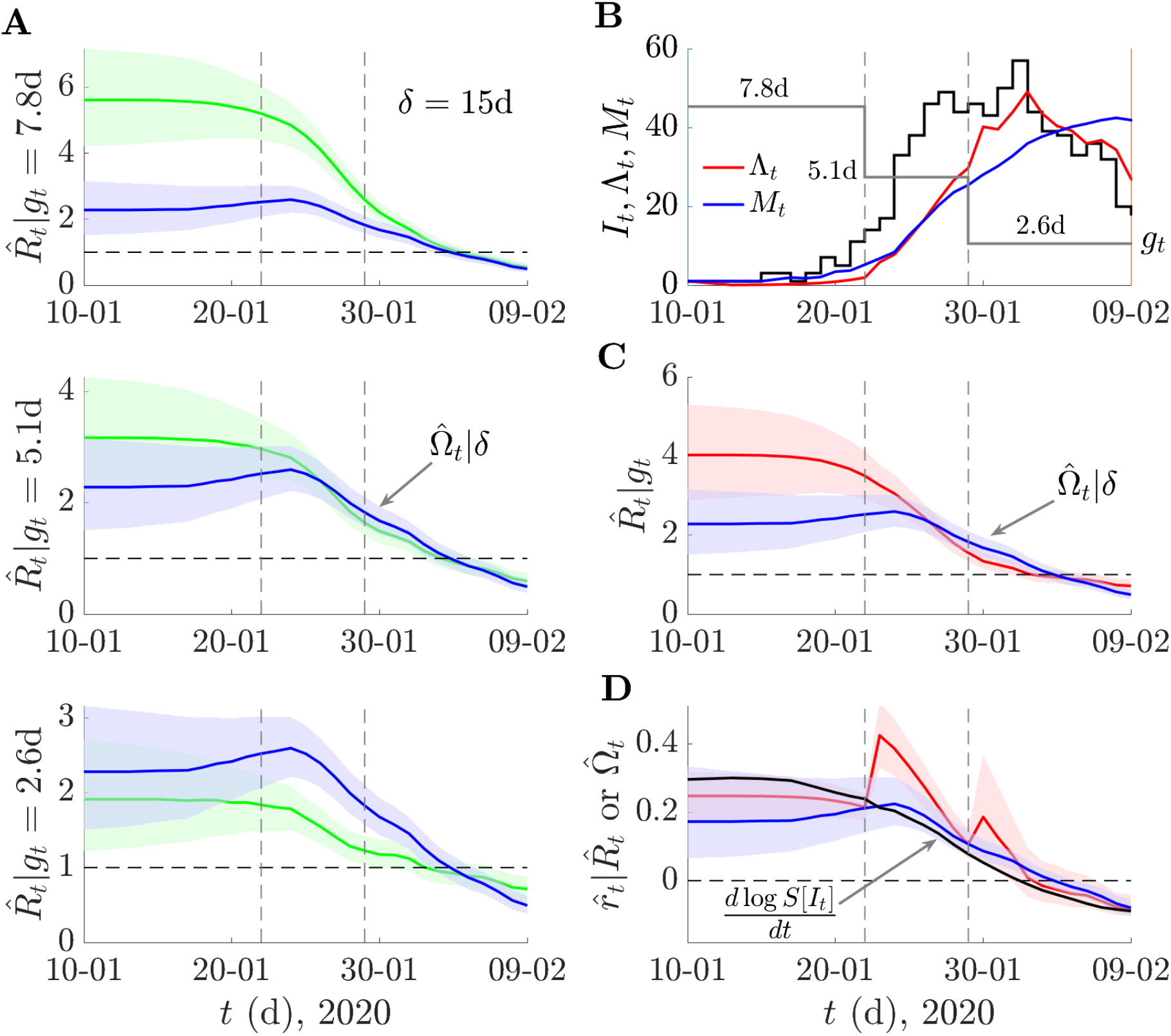
COVID-19 transmissibility in China under non-stationary generation times. We analyse COVID-19 data from [15], which spans 9^th^ January 2020 to 13 February 2020 and is known to feature a serial interval distribution that shortened in mean substantially from 7.8d to 2.6d (change times are shown as grey vertical lines). We assume that the serial interval approximates the generation time well and replicate the analysis from Figure 2 of [15]. In panel A, we compare estimates (green) of effective reproduction numbers, *R*, using fixed generation time distributions inferred in [15] (specified by their means *g*) against those of our angular reproduction number Ω (blue). We use EpiFilter [25] to obtain all estimates (means shown with 95% credible intervals) and find relative trends similar to those in Figure 2 of [15]. In panel B we plot the incidence (black) and the denominators we use to compute an *R* that does account for the generation time changes (Λ, red) and for Ω (*M*, blue). This *R* uses the different distributions inferred at the grey vertical change times (their means are in panel B and are also the fixed distributions of panel A in sequence). We plot these *R* and Ω estimates in panel C. In panel D we show the growth rates that are inferred from the *R* and Ω estimates of C (red and blue respectively) against that obtained from taking the smoothed log derivative (black).

We see that Ω provides a lower assessment of the initial transmissibility as compared to the *R* that is best informed by the changing *w* but that both agree in general and in particular at the important threshold between super- and subcritical spread. Interestingly, Ω indicates no sharp changes at the *w* change-times. This follows because the incidence is too small for those changes to shape overall transmissibility and matches the gradual *w* changes originally inferred in [15]. The distributions used in **Figure 6** provide a piecewise approximation to these variations. We also compare *r* estimates derived from *R* (red, from [14]), Ω (blue, from **Eq. (5)**) and the empirical log gradient of smoothed incidence 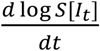. We find that the *r* from Ω agrees more closely with the empirical growth rate than the *r* from *R*, which somewhat by design shows jumps at the *w* change-points. While this analysis is not meant as a detailed study of COVID-19 in China, it does demonstrate the practical usefulness of Ω.

## Discussion

Quantifying the time-varying transmissibility of a pathogen remains an enduring challenge in infectious disease epidemiology. Changes in transmissibility may signify shifts in the dynamics of an epidemic of relevance to both preparedness and policymaking. While this challenge has been longstanding, the statistics that we use to summarise transmissibility have evolved from dispersibility [39] and incidence to prevalence ratios [40] to cohort [41] and instantaneous [22] reproduction numbers. While the last, which we have denoted *R*, has become the predominant metric of transmissibility, all of these proposed statistics ultimately involve a ratio between new infections and a measure of active infections (i.e., the denominator). Deciding on appropriate denominators necessitates some notion (implicit or explicit) of a generation time [42].

Difficulties in characterising these generation times and their changes substantially bias [6] estimates of transmissibility and have motivated recent works to propose the instantaneous growth rate, *r*, as a more reliable approach for inferring pathogen spread [20]. However, on its own, *r* is insufficient to resolve many of the transmission questions that *R* can answer and its computation may employ smoothing assumptions that are in some instances equivalent to the generation time ones behind *R* [4]. We formulated the novel angular reproduction number Ω, to merge some advantages from both *R* and *r* and to contribute to a more comprehensive view of transmissibility. By applying basic vector algebra (**Eqs. 1-3**), we encoded both changes to *R* and the generation time distribution, *w*, into a single time-varying metric, deriving Ω.

We found that Ω maintains the threshold properties and individual-level interpretability of *R* but responds to variations in *w*, in a manner consistent with *r* (**Figure 2**). Moreover, Ω indicates variations in transmissibility caused by *R* and *w* without requiring measurement of generation times (**Figure 3**). This is a consequence of its denominator, which is the root mean square of infections over a user-specified window δ that is relatively simple to tune (see Methods). We can interpret Ω = *a* > 1 as indicating that infections across δ need to be reduced by *a*^-1^. This reduces mean and root mean square infections by *a*^-1^ and causes Ω to equal 1. Further, Ω circumvents identifiability issues surrounding the joint inference or *R* and *w* [43] by refocussing on estimating the net changes produced by both. This improves our ability to *explain* the shifts in transmissibility underpinning observed epidemic dynamics and means Ω is essentially a reproduction number that provides individual-level interpretation of growth rates (**Eqs. 4-5**).

The benefits of this *r*-Ω correspondence are twofold. First, as interventions may alter *R, w* or *R* and *w* concurrently [15,18] situations can arise where *r* and *R* disagree across time on both the drivers and magnitude of transmissibility. While it may seem possible to minimise this issue by constructing alternative threshold statistics by directly combining *r* with assumed generation time structures, we find these statistics often exhibit worse performance and larger bias than Ω (**Supplementary Information**). Second, this disagreement can also occur when comparing pathogenic variants or epidemics (e.g., from diverse spatial or sociodemographic groups) with different but known and unchanging *w*. This study appears to be among the earliest to highlight these discrepancies, which can occur in multiple settings (see **Supplementary Information**). Realistic transmission landscapes possess all of the above complexities, meaning that relying solely on conventional measures of relative transmissibility can lead to contradictions.

We found that Ω consistently orders epidemics by growth rate while capturing notions of the average new infections per past infection (**Figure 4**). This suggests Ω blends advantages from *R* and *r*, with clearer assumptions (choice of window δ). However, Ω offers no advantage if we want to *predict* epidemic dynamics (see [35] for more on prediction-explanation distinctions). For this problem even an *R* inferred using a misspecified denominator performs equally well (**Figure 5**). This follows as only the product of any reproduction number and its denominator matter when determining the next incidence value. Iterations of this product underpin multi-step ahead predictions [38]. This may explain why autoregressive models, which ignore some characteristics of *w*, can serve as useful predictive models [44]. Other instances where Ω will not improve analysis are at times earlier than δ (due to edge effects [9]) and in periods of near zero incidence (there is no information to infer *R* either [24]). We summarised and compared key properties of *R, r* and Ω in **Table *1***.

There are several limitations to our study. First, we only examined biases inherent to *R* due to the difficulty of measuring the generation time accurately and across time. While this is a major limitation of existing transmissibility metrics [15], practical surveillance data are also subject to under-reporting and delays, which can severely diminish the quality of any transmissibility estimates [23,43,45]. While Ω ameliorates issues due to generation time mismatch, it is as susceptible as *R* and *r* to surveillance biases and corrective algorithms (e.g., deconvolution methods [46]) should be applied before inferring Ω. Second, our analysis depends on renewal and compartmental epidemic models [22]. These assume random mixing and cannot account for realistic contact patterns. Despite this key structural uncertainty, there is evidence that well-mixed and network models are comparable when estimating transmissibility [47].

Although the above limitations can, in some instances, reduce the added value of improving the statistics summarising transmissibility, we believe that Ω will be of practical and theoretical benefit, offering complementary insights to *R* and *r* and forming part of a more comprehensive epidemic analytic toolkit. Its similarity in formulation to *R* means it is as easy to compute using existing software and therefore can be deployed on dashboards and updated in real time to improve situational awareness. Further, Ω improves comparison and communication of the relative risks of circulating variants or epidemics among diverse groups, avoiding *R*-*r* contradictions provided the known parameter, δ, is fixed. This supplements *R*, which is hard to contextualise [20] when *w* is misspecified or varying and hence compare across groups, as each group may have distinct and correspondingly poorly specified denominators. Last, Ω can help probe analytical questions about how changes in *R* and *w* interact because it presents a common framework for testing how variations in either influence overall transmissibility.

## Methods

### Inferring angular reproduction numbers across time

We outline how to estimate Ω_*t*_ given a time series of incident infections 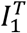, with *T* defining the present or last available data timepoint i.e., 1 ≤*t* ≤*T*. Because Ω_*t*_ simply replaces the total infectiousness Λ_*t*_, used for computing *R*_*t*_, with the root mean square of the new infection time series (see **Figure 1**), *M*_*t*_, we can obtain Ω_*t*_ from standard *R*_*t*_ estimation packages with minor changes. This requires evaluating *M*_*t*_ over some user-defined, backward sliding window of size δ. Under a Poisson (Pois) renewal model this follows as in **Eq. (6)** for timepoint *t*.

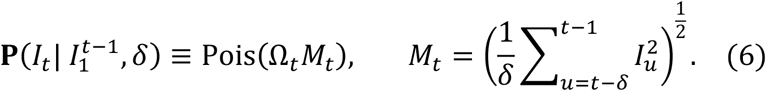

The choice of δ is mostly arbitrary but should be sufficiently long to capture most of the likely probability mass of the unknown generation time but not overly long since it induces an edge effect (similar to the windows in [9,37]). We found a suitable heuristic to be twice or thrice the initial expected mean generation time (*g*_0_). We can then input *M*_*t*_ and *I*_*t*_ into packages such as EpiEstim [9] or EpiFilter [25] to estimate Ω_*t*_ with 95% credible intervals.

Due to the similarity between computing *R*_*t*_ and Ω_*t*_ we only specify the latter but highlight that replacing *M*_*t*_ with Λ_*t*_ yields the expressions for evaluating any equivalent quantities from *R*_*t*_. The only difference relates to how the growth rates *r*_*t*_ are computed. We estimate *r*_*t*_ from *R*_*t*_ by applying the generation time, 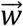, based transformation from [14]. For a correctly specified 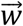 this gives the same result as the smoothed derivative of the incidence curve [4]. We derive *r*_*t*_ from Ω_*t*_ using **Eq. (5)**, which follows from rearranging **Eq. (4)** into 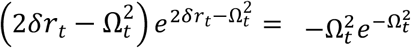. This expression then admits Lambert W function solutions. In all estimates of *r*_*t*_ we propagate uncertainty from the posterior distributions (see below) over *R*_*t*_ or Ω_*t*_.

We applied EpiFilter in this study due to its improved extraction of information from 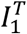. This method assumes a random walk state model for our transmissibility metric as in **Eq. (7)** with ϵ_*t*−1_ as a normally distributed (Norm) noise term and η as a free parameter (default 0.1).

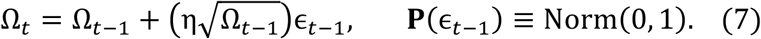

The EpiFilter approach utilises Bayesian smoothing algorithms incorporating the models of **Eq. (6)-(7)** and outputs the complete posterior distribution 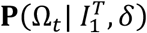 with *T* as the complete length of all available data (i.e., 1 ≤*t* ≤*T*). We compute our mean estimates 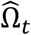 and 95% credible intervals from this posterior distribution and these underlie our plots in **Figures 3-4**.

EpiFilter also outputs the one-step-ahead predictive distributions 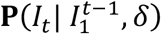, which we use in **Figure 5**. There we quantify predictive accuracy using the predicted mean square error PMSE and the accumulated prediction error APE, defined as in **Eq. (8)** [36,37] with 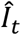 as the posterior mean estimate from 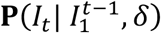 and 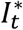 as the true simulated incidence. These are computed with 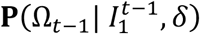 and not 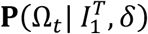, ensuring no future information is used.

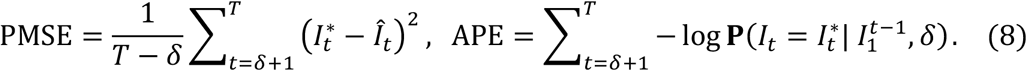

We collectively refer to these as distance metrics 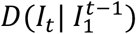 and construct their distributions, **P**(*D*), over many replicates of simulated epidemics. Last, we use 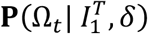 to compute the posterior distribution of the growth rate 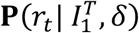 and hence its estimates as in **Eq. (5)**. More details on the EpiFilter algorithms are available at [25,31,48]. We supply open-source code to reproduce all analyses at https://github.com/kpzoo/Omega as well as functions in MATLAB and R to allow users to estimate Ω_*t*_ from their own data.

## Data availability statement

All data and code underlying the analyses and figures of this work are freely available (in R and MATLAB) at: https://github.com/kpzoo/Omega

