## supplementary information for "Angular reproduction numbers improve estimates of transmissibility when disease generation times are misspecified or time-varying"


### Deriving threshold statistics from growth rate estimates

In the main text we derived the angular reproduction number, $\Omega$, which responds to variations in transmissibility caused by changes to either or both of the effective reproduction number *R*, and the generation time distribution *w*. An interesting property of $\Omega$ is that it is consistent with the time-varying growth rate *r* (unlike *R*), but still possesses a useful individual-level threshold interpretation (unlike *r*). However, there are alternative ways to convert *r* into a threshold-like statistic about 1 [1]. Specifically, we may apply any of the transformations from [2], which relate *R* to *r* under a mean generation time, but replace the generation time with a window parameter $\alpha$ that we can customise. These transformations all guarantee (at least for a constant true *R*) that the sign of *r* is consistent with that of $R(\alpha)$-1, with $R(\alpha)$ as the threshold statistic.

We investigate the most commonly used transformations in **Eq. (S1)**. The left-side expression assumes a gamma distributed generation time structure with shape $\sigma$ and scale $\alpha\sigma^{-1}$ (this has mean $\alpha$). The right-side relation considers a normally distributed structure with mean $\alpha$ and standard deviation $\sigma$. These equations are taken from [2].

$$R(\alpha) = \left(1 + \frac{r\alpha}{\sigma}\right)^{\sigma}, \qquad R(\alpha) = e^{r\alpha - \frac{1}{2}r^2\sigma^2}. \quad (\mathrm{S1})$$

For an exponentially distributed generation time $\sigma$=1 on the left-side equation yielding $R(\alpha) = 1 + r\alpha$. At the other extreme of a deterministic or fixed generation time distribution $\sigma$=0 on the right-side expression (or $\sigma$ becomes infinite on the left-side equation), resulting in $R(\alpha) = e^{r\alpha}$.

We investigate if threshold statistics derived from these canonical relationships will outperform our proposed angular reproduction number. We repeat the simulations from **Figure 3** of the main text, which features step-change and cyclic dynamics for Ebola virus disease (EVD) and COVID-19 with time-varying in mean generation times. We present mean estimates of $\Omega$ and $r$ from these simulations in **Figure S1**. The latter is computed as a smoothed log derivative of incidence and demonstrates one issue with these alternate metrics. Because $R(\alpha)$ relies on a model-agnostic estimate of $r$, it is sensitive to smoothing assumptions [3], which are non-trivial to benchmark in the absence of epidemiological knowledge. This can limit our ability to signal any changes in the generation time (note that these changes are only inferable when incidence is sufficiently large [4]). We observe this in the bottom panel of **Figure S1** where $\Omega$ spikes in response to a contraction of the generation time but $r$ shows no discernible deviation.

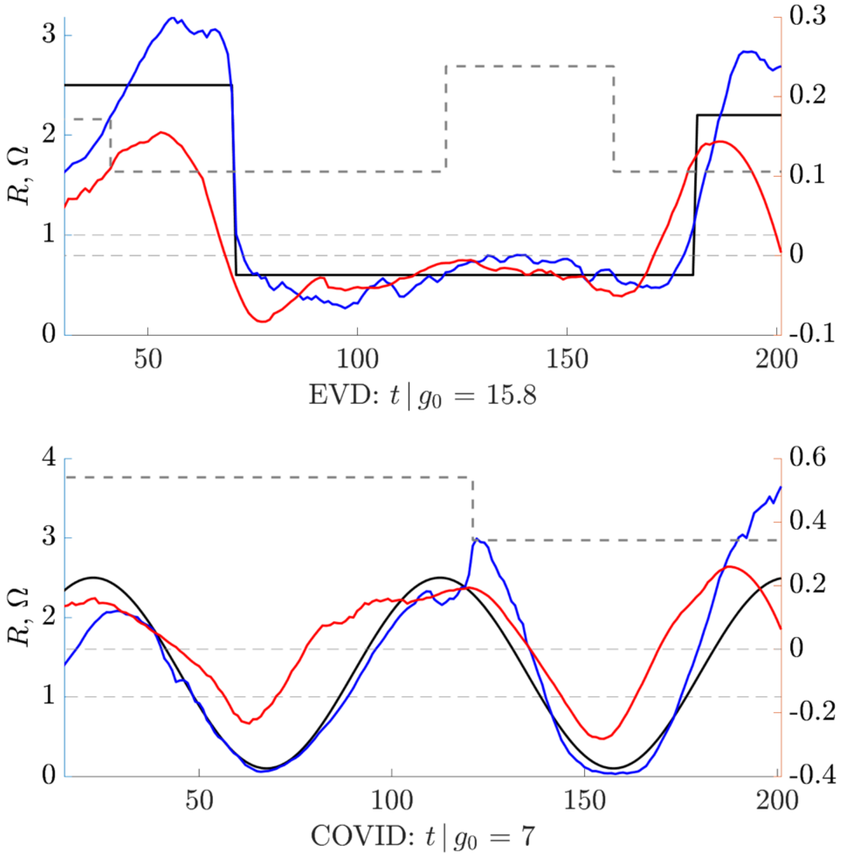


**Figure S1: Mean measures of transmissibility.** We repeat the simulations in **Figure 3** from the main text under gamma distributed generation times with changing mean (grey dashed, with $g_0$ as the initial mean). We show posterior mean estimates of the angular reproduction number $\Omega$ (blue, derived using EpiFilter [5]) and the true reproduction number $R$ (black) on left y-axes. We overlay estimates of the growth rate $r$ (red, computed as the log derivative of the smoothed incidence using Savitzky-Golay filters [6]) on right y-axes. Crossing times between $r$ and 0 and $R$ or $\Omega$ and 1 should delineate between subcritical and supercritical transmission.

We then compute $R(\alpha)$ under exponential, deterministic, gamma and normal generation time distribution assumptions as described above. We input our $r$ estimates into **Eq. (S1)** for various window values $\alpha$ and set $\delta=2\alpha$. We chose this relationship between $\alpha$ and $\delta$ as the first is a proxy for a distribution mean, while the second is a proxy for its support. **Figures S2** and **S3** present the threshold statistics and $\Omega$ for EVD and COVID for these window or free parameter choices on simulated data from **Figure 3** of the main text. We find the threshold statistics are less robust to changes in window choices than the angular reproduction number and become unreliable even when $\alpha$ closely approximates the original mean generation time $g_0$. The choice of formulation of $R(\alpha)$ also strongly determines the properties of the resulting statistic.

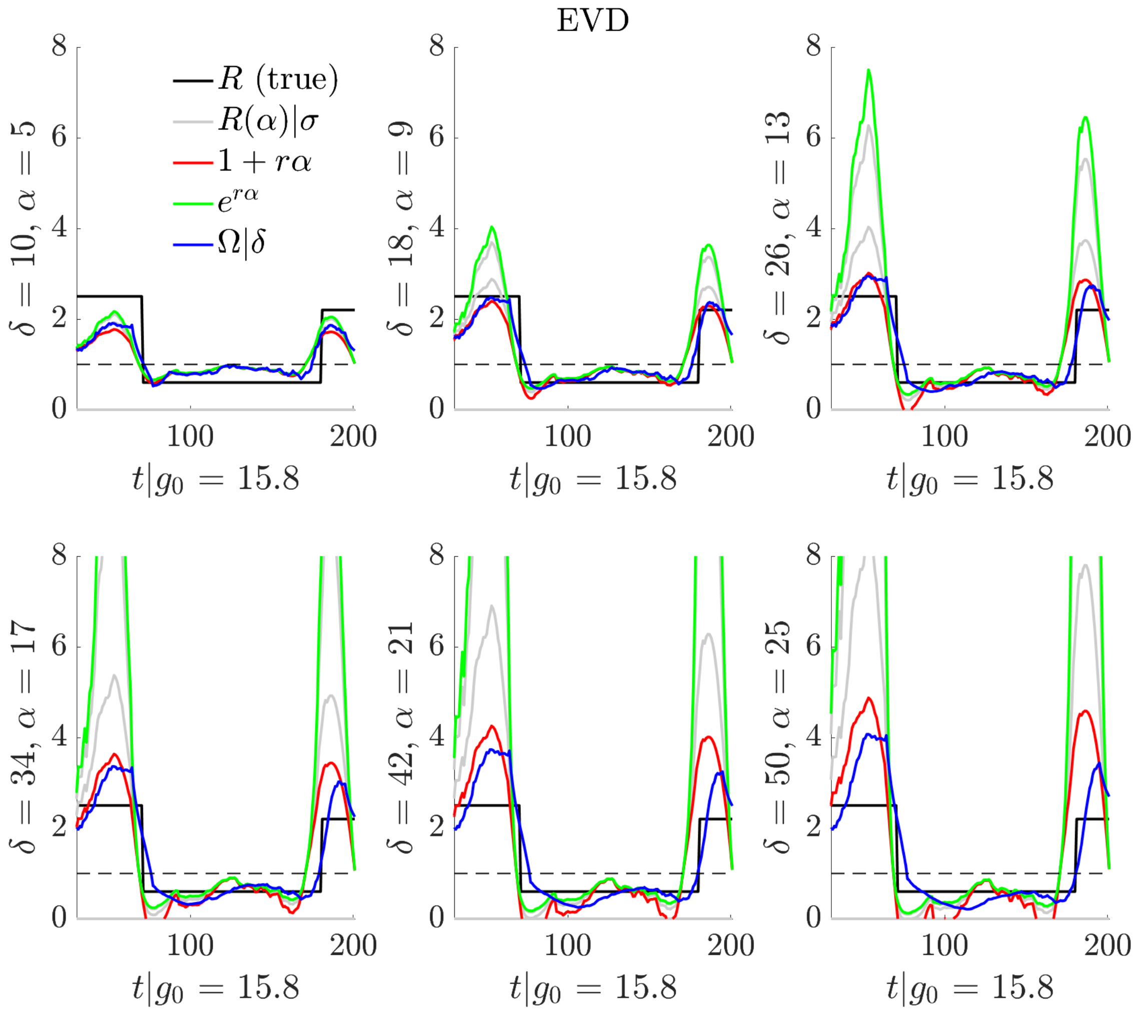


**Figure S2: Relationships among transmissibility metrics for EVD.** We explore threshold statistics computed using **Eq. (S1)** under structural assumptions from [2] and compare against the angular reproduction number Ω (blue) at various window sizes of α and $\delta$=2α respectively. These EVD simulations are from **Figure 3** of the main text with the true reproduction number (black) undergoing step dynamics. The threshold *R*(α) statistics combine mean growth rate estimates from **Figure S1** with α, which serves as a proxy for the mean generation time under structural assumptions of exponential (red) and deterministic (green) distributions as well as intermediary normal and gamma distributions (grey, with additional spread parameter $\sigma$).

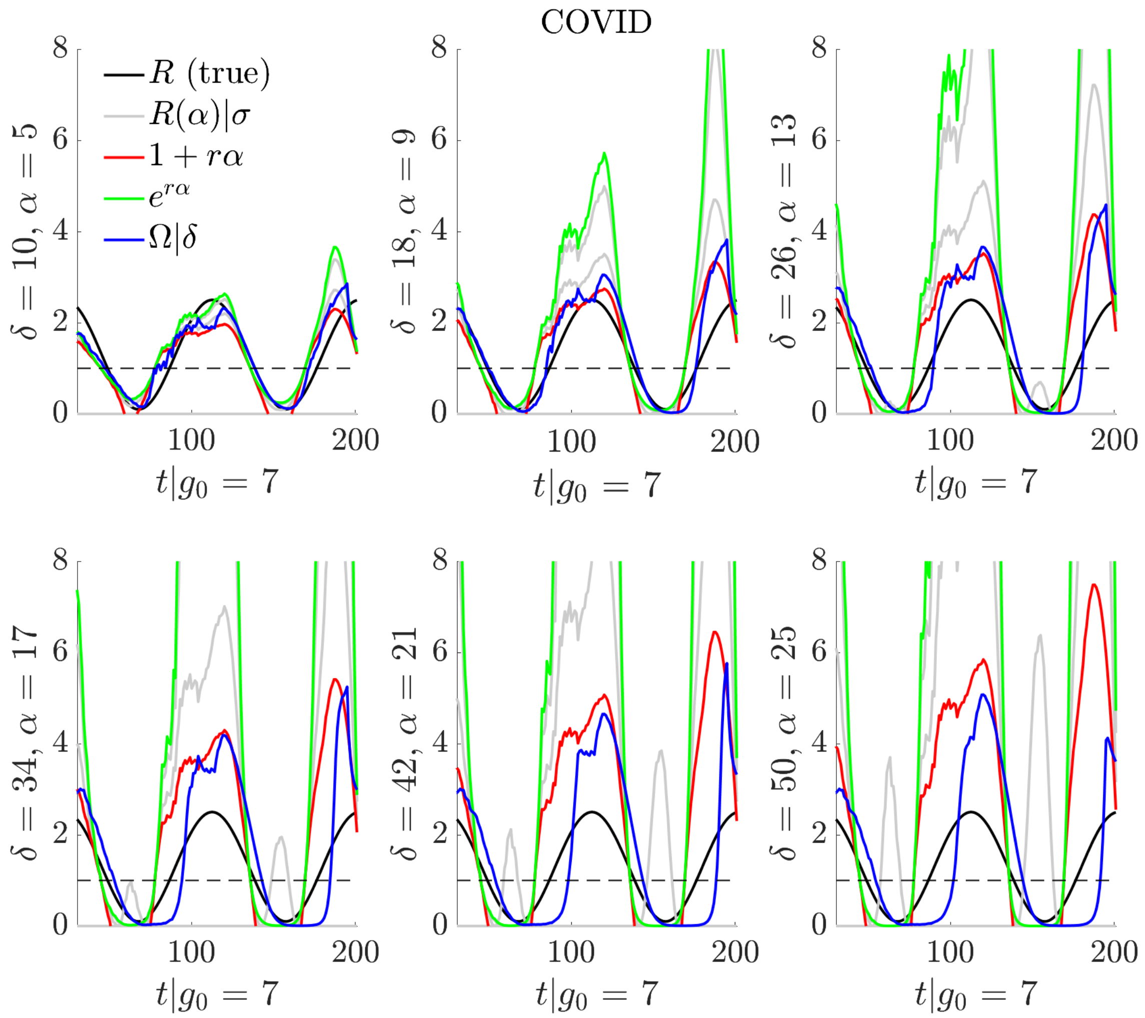


**Figure S3: Relationships among transmissibility metrics for COVID.** We investigate the threshold statistics *R*(α) computed in **Eq. (S1)** under the structural assumptions from [2] and compare against the angular reproduction number Ω (blue) at various window sizes of α and $\delta$=2α respectively. These COVID simulations are from **Figure 3** of the main text with cyclical true reproduction numbers (black). The threshold statistics combine mean estimates of growth rates from **Figure S1** with α, which acts as a proxy for the mean generation time under the structural assumptions of exponential (red), deterministic (green) and intermediary normal and gamma distributions (which require an additional spread parameter $\sigma$ and are in grey).

We observe reasonable correspondence among the arbitrary threshold statistics of $R(\alpha)$ and $\Omega$ at small window sizes but substantial discordance as those windows become larger. This discordance even persists for some $\alpha$ that are close to the true mean generation time. Across many scenarios the $R(\alpha)$ statistics appreciably overestimate transmissibility and appear to be biased in determining the transition from subcritical to supercritical transmission. Importantly, $\Omega$, which makes fewer assumptions than $R(\alpha)$, is simpler to tune as it varies less appreciably than $R(\alpha)$ when its window is incorrectly specified, and it more accurately signals transitions. Note that under very large windows all statistics, including $\Omega$, do not perform well. This results from those windows causing extensive over-smoothing and is a problem even when inferring *R* for a disease with stationary and perfectly known generation time as shown in [7,8].

### Understanding the inversion of growth rate and reproduction number rankings

In **Figure 4** of the main text, we demonstrated that it is possible for co-circulating epidemics or variants to possess inverted rankings on their reproduction numbers and growth rates due to the impact of transmissibility changes caused by an intervention (or any related effect). This phenomenon means that pre-intervention a variant may have the larger *R* but smaller *r*, then post-intervention it may instead possess both the larger *R* and *r*. Intervention relaxations may conversely cause a variant with both smaller *R* and *r* to have the larger *r* post-relaxation. This phenomenon, as far as we are aware, has not been described. Here we provide mathematical arguments to show that inversion can be prevalent and is not simply an artefact of **Figure 4**.

We consider two epidemics or variants with reproduction numbers and mean generation times related as $R_1 = \alpha R_2$ and $g_1 = \beta g_2$. Assume $\beta > \alpha > 1$ so $R_1 > R_2$ and $g_1 > g_2$. We define an intervention as a multiplicative change by factor $0 < \eta < 1$ so that post-intervention the $j^{\text{th}}$ variant has reproduction number $R_{j\eta} = \eta R_j$. This intervention hence maintains the ordering $R_{1\eta} > R_{2\eta}$. An inversion occurs if growth rates before and after the intervention are such $r_1 < r_2$ but $r_{1\eta} > r_{2\eta}$. We test this possibility under the fundamental exponential and deterministic generation time distributions, which also are the extremes of gamma and normally distributed generation times (see **Eq. (S1)**). In this framework, a relaxation causes a multiplicative change $\eta > 1$ and can lead to different inversions that we also outline below.

For the exponential case $r_j = g_j^{-1}(R_j - 1)$. We find $r_1 < r_2$ when $\beta^{-1}(\alpha R_2 - 1) < R_2 - 1$. This implies we need $R_2(\beta - \alpha) > \beta - 1$ and hence $R_2 > (\beta - 1)(\beta - \alpha)^{-1}$. We can satisfy the post-intervention inequality $r_{1\eta} > r_{2\eta}$ if $\beta^{-1}(\alpha\eta R_2 - 1) > \eta R_2 - 1$. The algebra is similar here and the condition for inversion is that $\eta R_2 < (\beta - 1)(\beta - \alpha)^{-1}$. In the deterministic case $r_j = g_j^{-1} \log R_j$ so $r_1 < r_2$ implies $\beta^{-1} \log \alpha R_2 < \log R_2$. This leads to $\log R_2 > \log \alpha^{\frac{1}{\beta-1}}$ and the condition $R_2 > \alpha^{\frac{1}{\beta-1}}$. Post-intervention $r_{1\eta} > r_{2\eta}$ occurs when $\beta^{-1} \log \alpha\eta R_2 > \log \eta R_2$. After some rearrangement this yields the analogous condition that $\eta R_2 < \alpha^{\frac{1}{\beta-1}}$. Consequently, whenever an intervention satisfies this pair of inequalities, we can expect an inversion.

As an example, let the second variant have 50% shorter mean generation time i.e., $\beta = 2$. We then need $R_2 > (2 - \alpha)^{-1}$ and $\eta R_2 < (2 - \alpha)^{-1}$ for the exponential case or $R_2 > \alpha$ and $\eta R_2 < \alpha$ for the deterministic one. If we set $R_1 = 1.5R_2$, these boundaries are 2 and 1.5 respectively. Many combinations of $R_2$ and $\eta < 1$ achieve inversion but the structure of the generation time is important for defining the parameter region of inversion. Similarly, when $r_1 > r_2$ pre-relaxation and $r_{1\eta} < r_{2\eta}$ post-relaxation we find that $R_2 < (\beta - 1)(\beta - \alpha)^{-1}$ and $\eta R_2 > (\beta - 1)(\beta - \alpha)^{-1}$ for exponential generation time distributions while $R_2 < \alpha^{\frac{1}{\beta-1}}$ and $\eta R_2 > \alpha^{\frac{1}{\beta-1}}$ for deterministic ones. Many $R_2$ and $\eta < 1$ pairs satisfy these conditions.

The intuition behind inversion is that changes in the reproduction numbers affect growth rates differently (if generation times are diverse) while also being constrained to always satisfy $R_t = 1 \Leftrightarrow r_t = 0$. Generally, it may be difficult (especially if surveillance is imperfect) to disentangle the impacts of interventions on reproduction numbers and generation times or to even track the structure of the generation time distribution. This motivates our proposed metric $\Omega_t$, which maintains $\Omega_t = 1 \Leftrightarrow r_t = 0$ and keeps the intuition of $R_t$ in terms of how much control effort we require but does not suffer any inversion (with respect to $r_t$).
